## Supplemental Information for "Call detail record aggregation methodology impacts infectious disease models informed by human mobility"

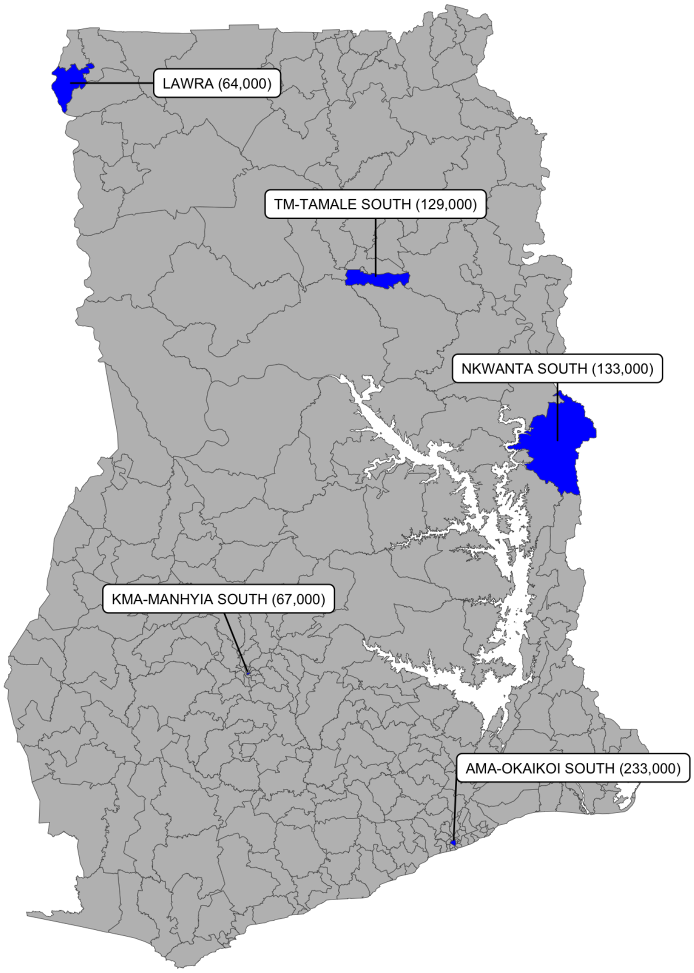


***Supplemental Figure 1. Epidemic Introduction locations.*** *To assess the sensitivity of the SIER model to the introduction location, we introduced disease into three urban districts: AMA-Okaikoi South, KMA-Manhyia South, TM-Tamale South and two rural districts Nkwanta South, and Lawra. Location labels show population size.*

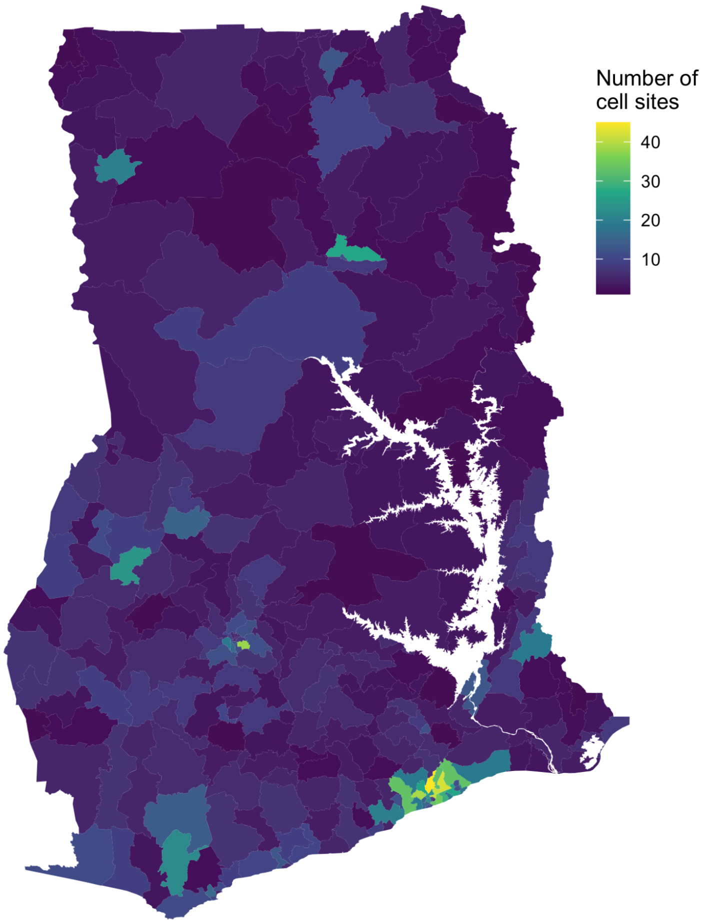


***Supplemental Figure 2. The number of cell sites per district.*** *The spatial distribution of cell sites, showing a high density of cell sites in urban areas.*

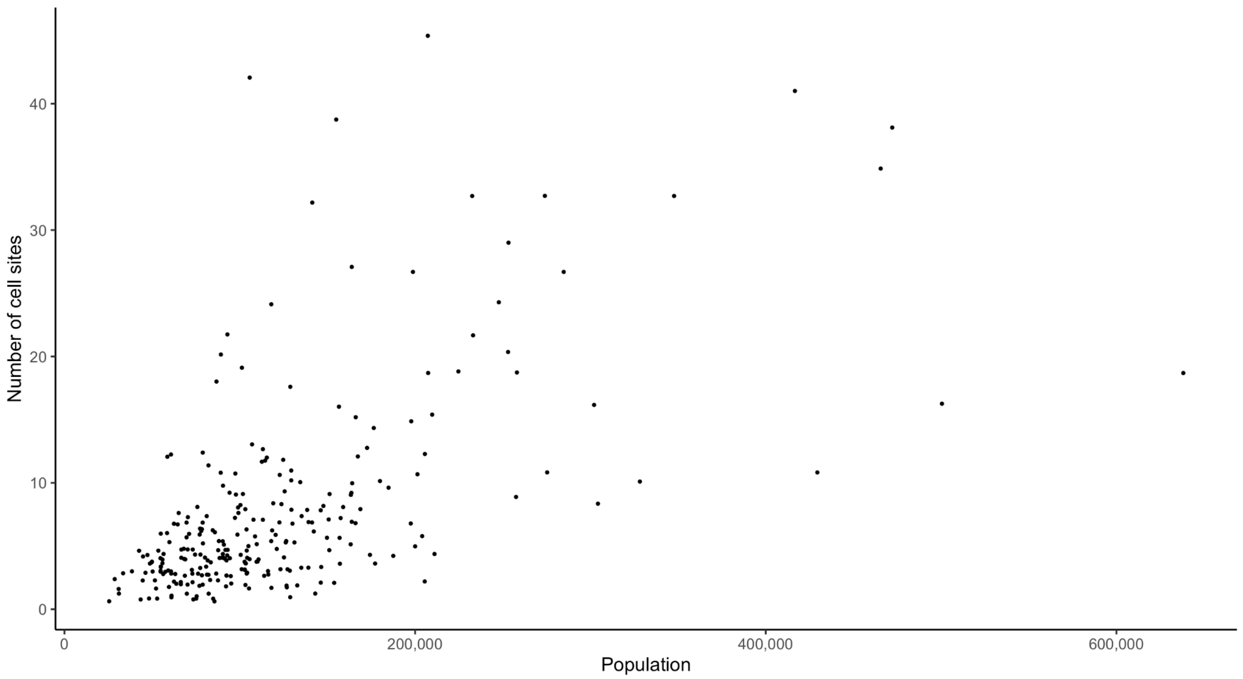


***Supplemental Figure 3. Number of cell sites by population.*** *The number of cell sites compared to the population in individual districts.*

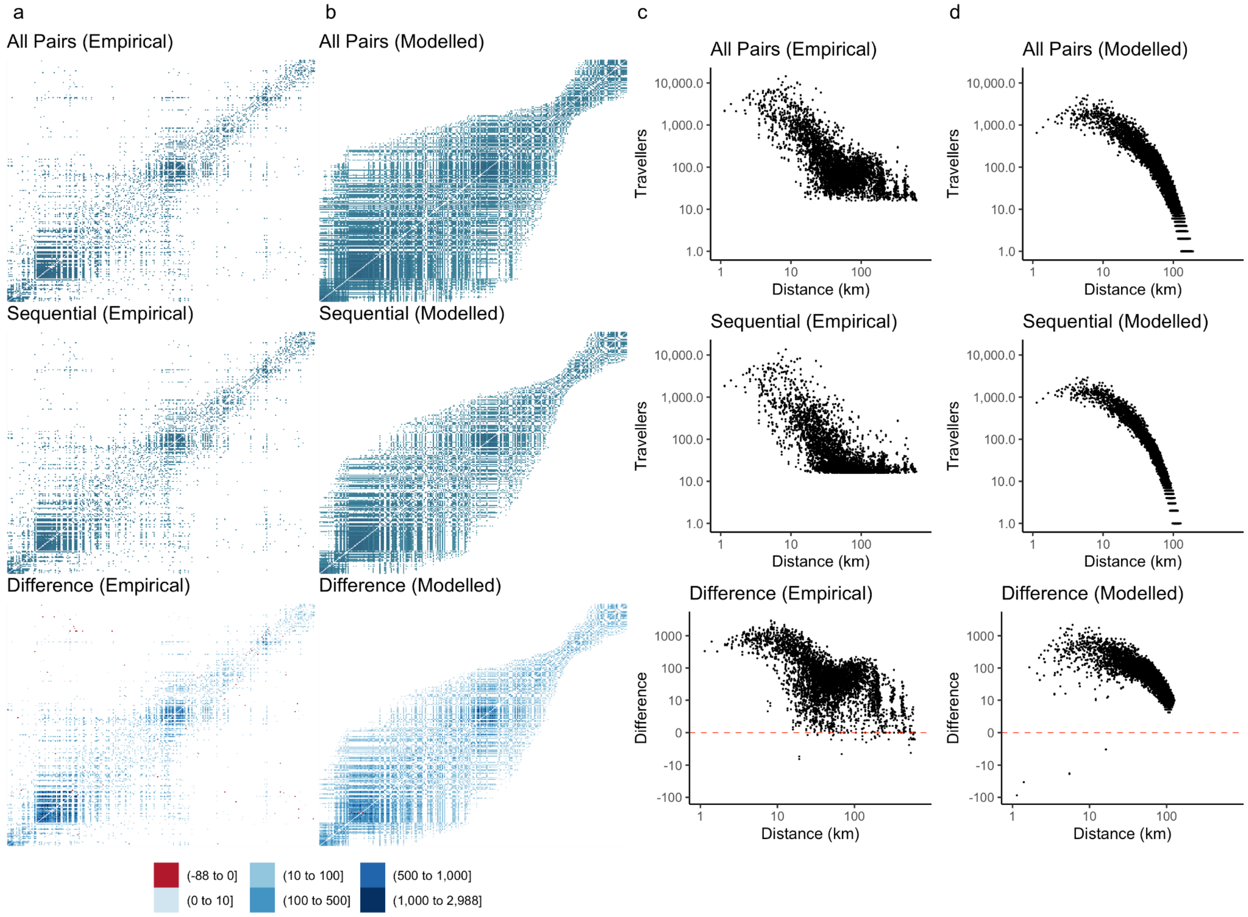


**Supplemental Figure 4*. Comparison of empirical and modelled travel networks.*** *a) Empirical networks from each aggregation methodology. b) Movement networks modelled using the exponential gravity model. Distance kernels show the number of travellers by the distance of network connections in the c) empirical and d) modelled networks.*

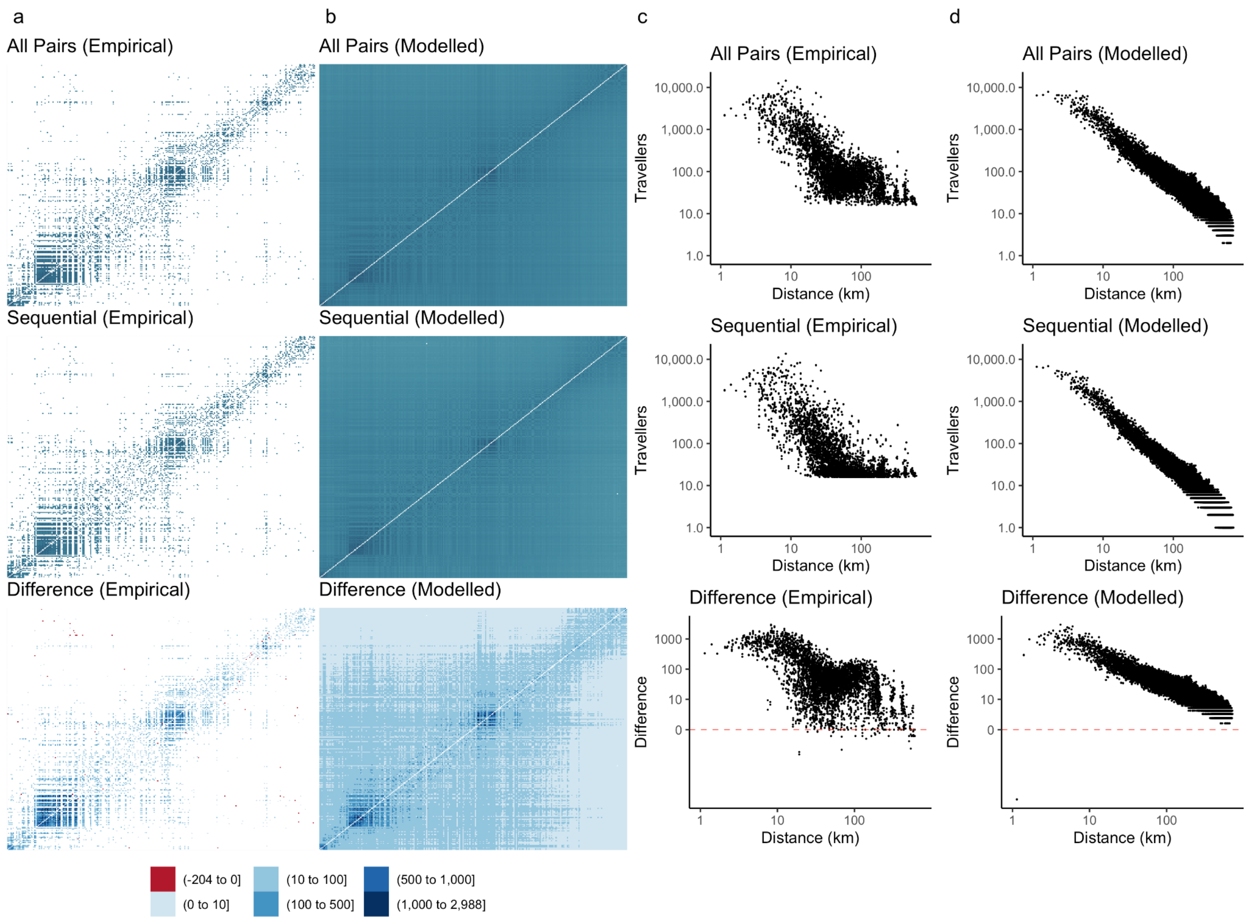


**Supplemental Figure 5*. Comparison of empirical and modelled travel networks.*** *a) Empirical networks from each aggregation methodology. b) Movement networks modelled using the power law gravity model. Distance kernels show the number of travellers by the distance of network connections in the c) empirical and d) modelled networks.*

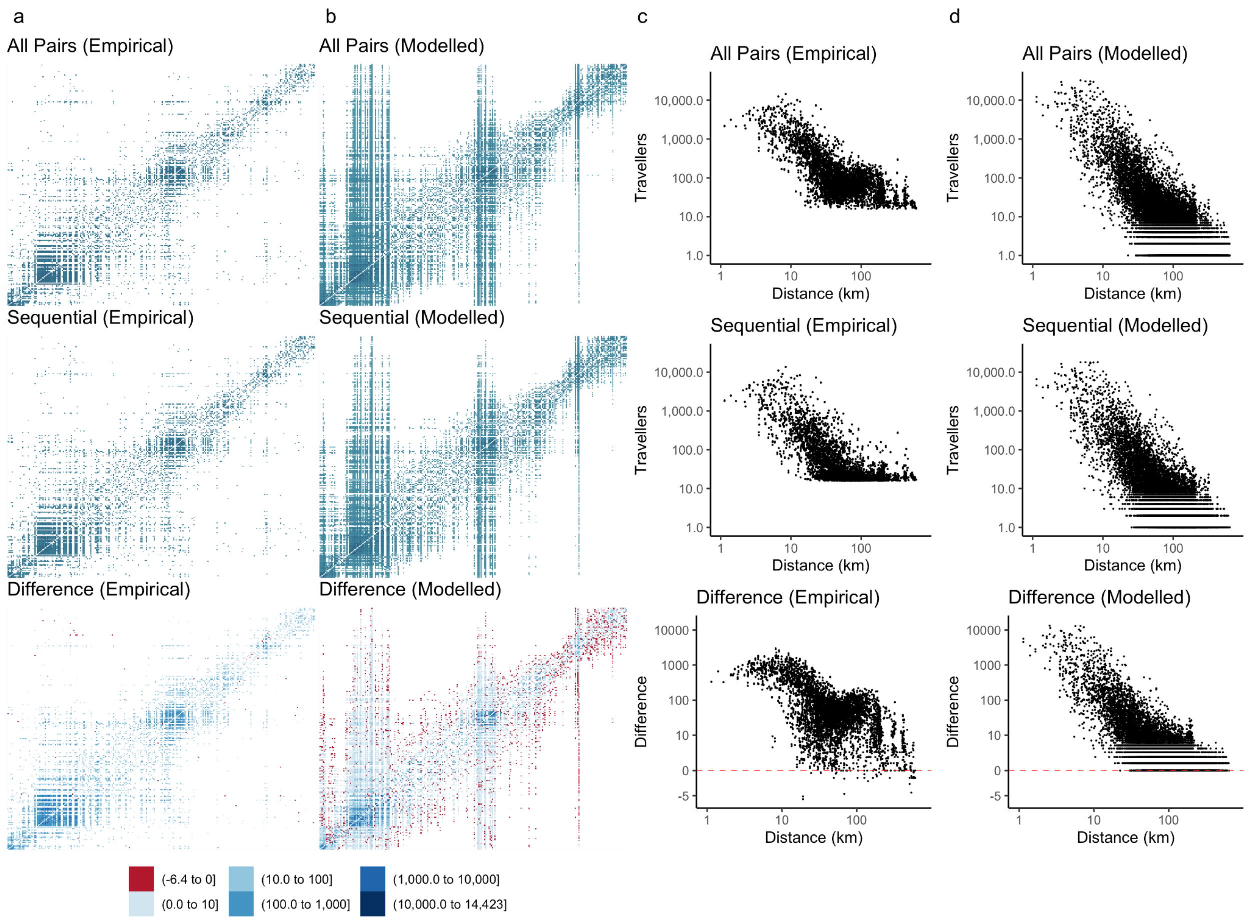


**Supplemental Figure 6*. Comparison of empirical and modelled travel networks.*** *a) Empirical networks from each aggregation methodology. b) Movement networks modelled using the radiation model. Distance kernels show the number of travellers by the distance of network connections in the c) empirical and d) modelled networks.*

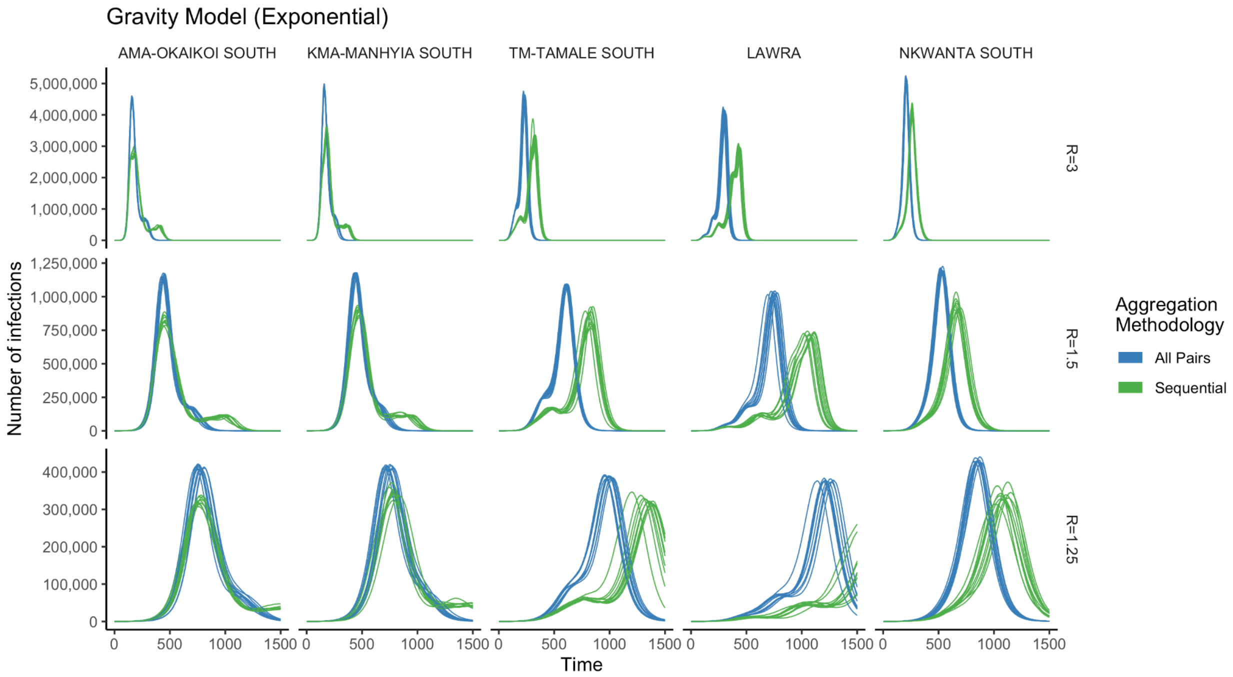


**Supplemental Figure 7*. Comparison of modelled national epidemics by aggregation methodology.*** *The difference in the national epidemic modelled for different introduction locations and different values of* $R_0$*. Epidemics were modelled 10 times for each combination of aggregation methodology, introduction location, and* $R_0$*. The first three districts (from left) are in urban centres of Accra, Kumasi, and Tamale, while the second two districts are rural districts in North West and East Ghana.*

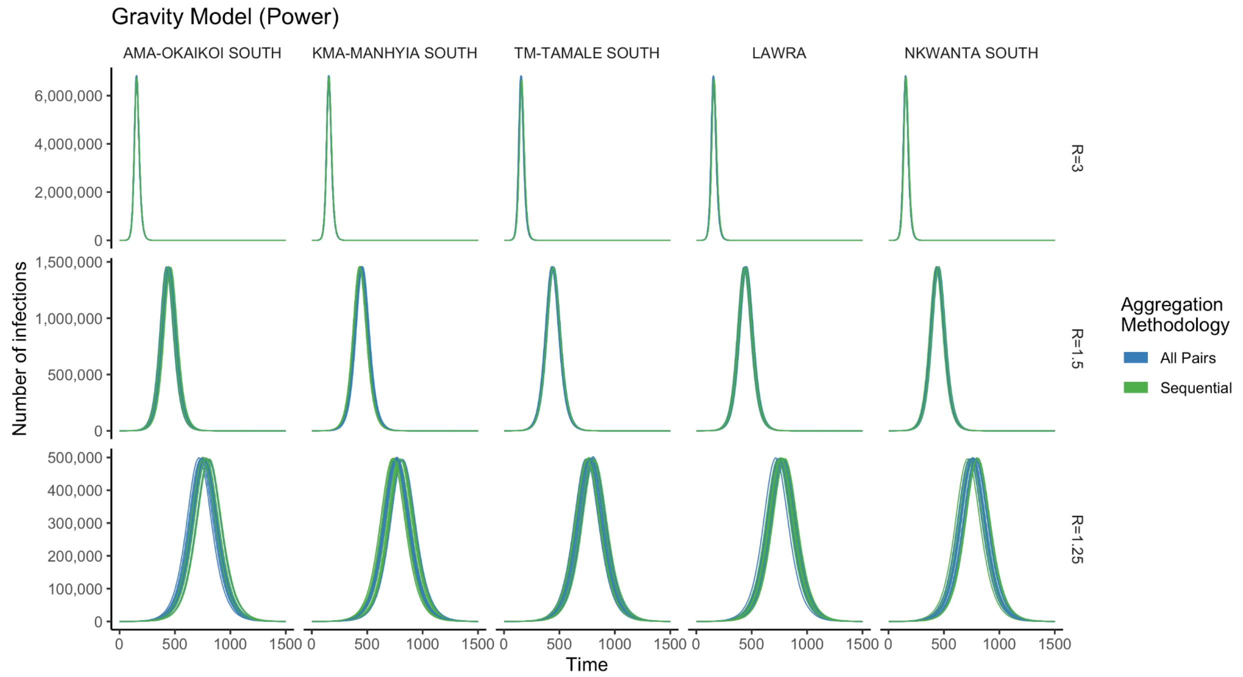


**Supplemental Figure 8*. Comparison of modelled national epidemics by aggregation methodology.*** *The difference in the national epidemic modelled for different introduction locations and different values of* $R_0$*. Epidemics were modelled 10 times for each combination of aggregation methodology, introduction location, and* $R_0$*. The first three districts (from left) are in urban centres of Accra, Kumasi, and Tamale, while the second two districts are rural districts in North West and East Ghana.*

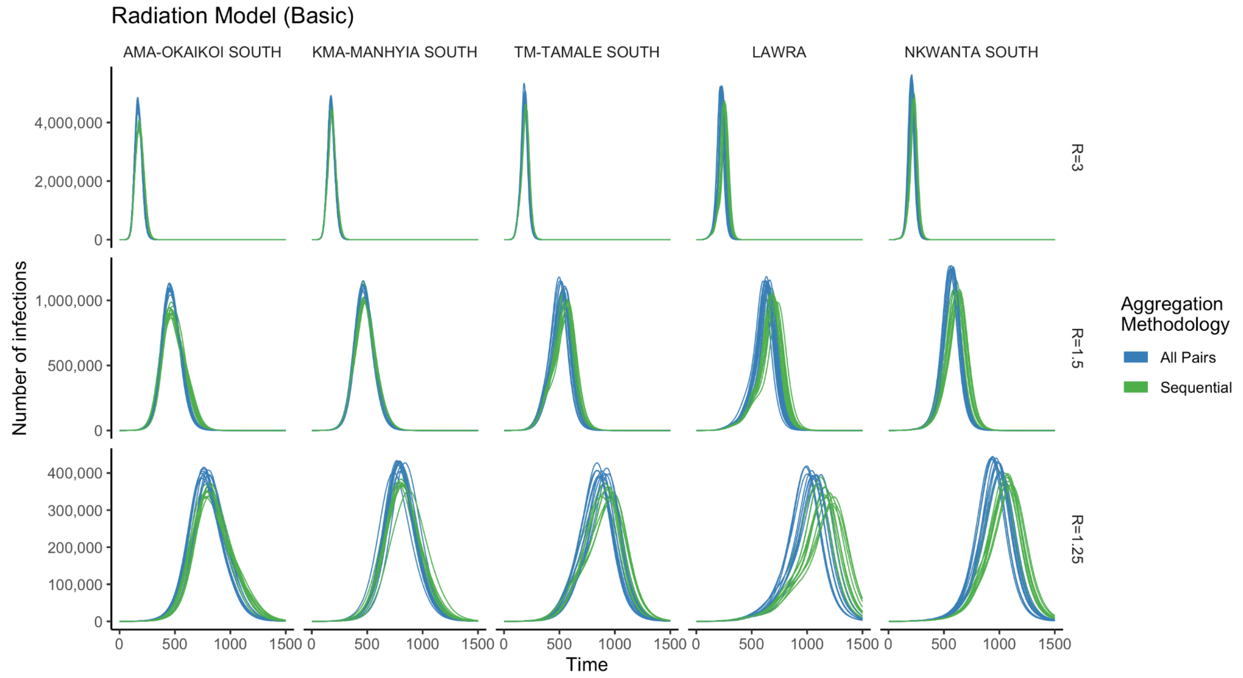


**Supplemental Figure 9*. Comparison of modelled national epidemics by aggregation methodology.*** *The difference in the national epidemic modelled for different introduction*

*locations and different values of $R_0$. Epidemics were modelled 10 times for each combination of aggregation methodology, introduction location, and $R_0$. The first three districts (from left) are in urban centres of Accra, Kumasi, and Tamale, while the second two districts are rural districts in North West and East Ghana.*

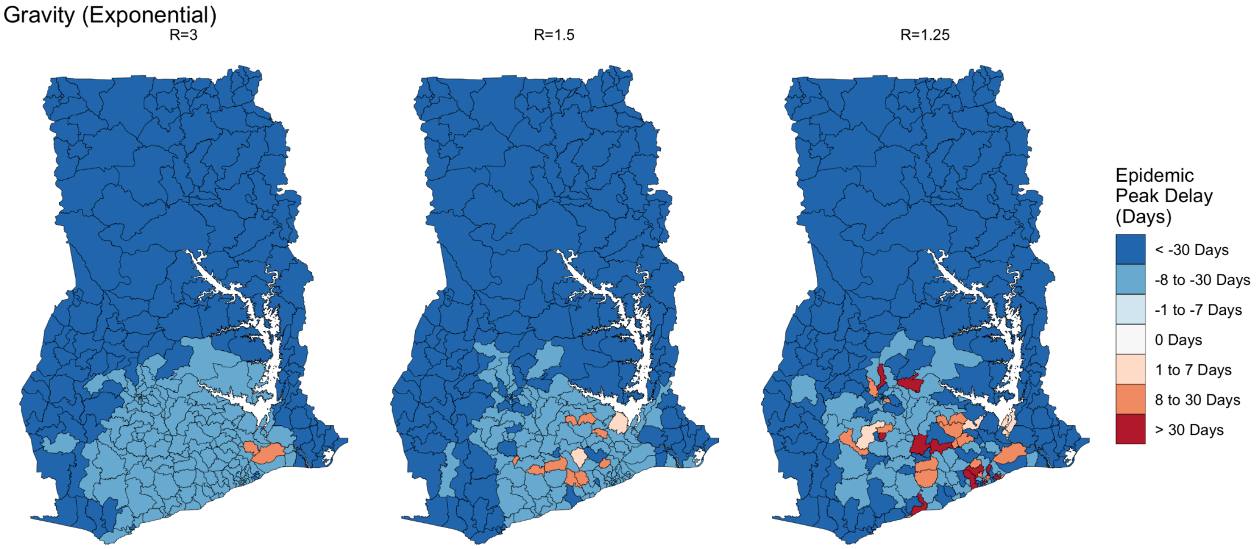


***Figure 10. Influence of introduction location and $R_0$ on the difference between aggregation methodologies.*** *Difference between the timing of the peak of a modelled epidemic informed my mobility data modelled using the exponential gravity model. Negative numbers indicate that the epidemic predicted based on the all pairs methodology was earlier than the epidemic predicted based on the sequential methodology.*

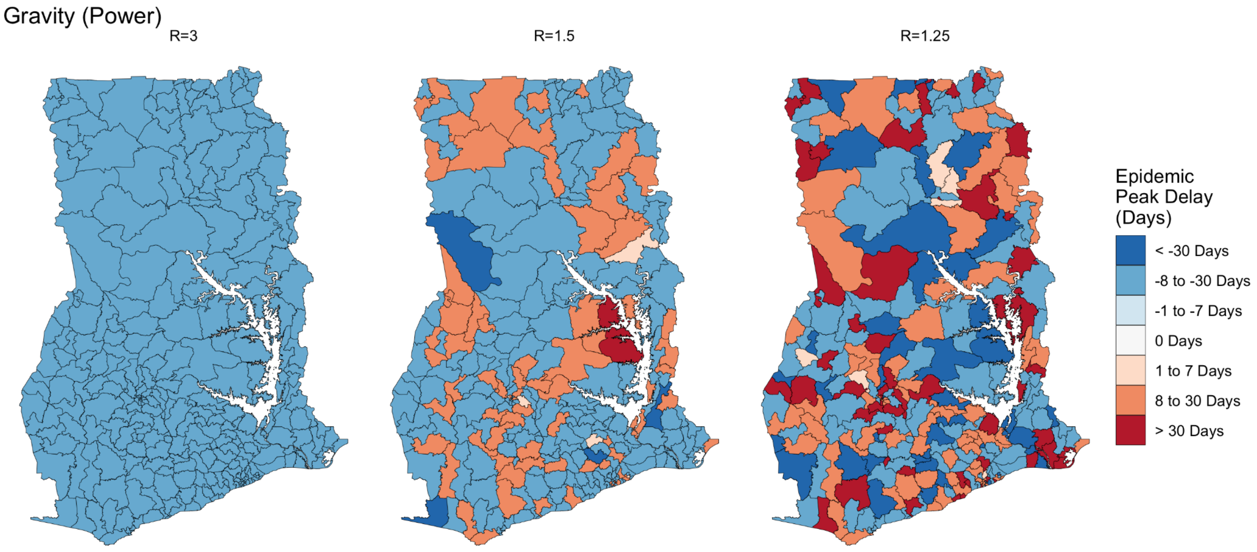


***Supplemental Figure 11. Influence of introduction location and $R_0$ on the difference between aggregation methodologies.*** *Difference between the timing of the peak of a*

*modelled epidemic informed my mobility data modelled using the power law gravity model. Negative numbers indicate that the epidemic predicted based on the all pairs methodology was earlier than the epidemic predicted based on the sequential methodology.*

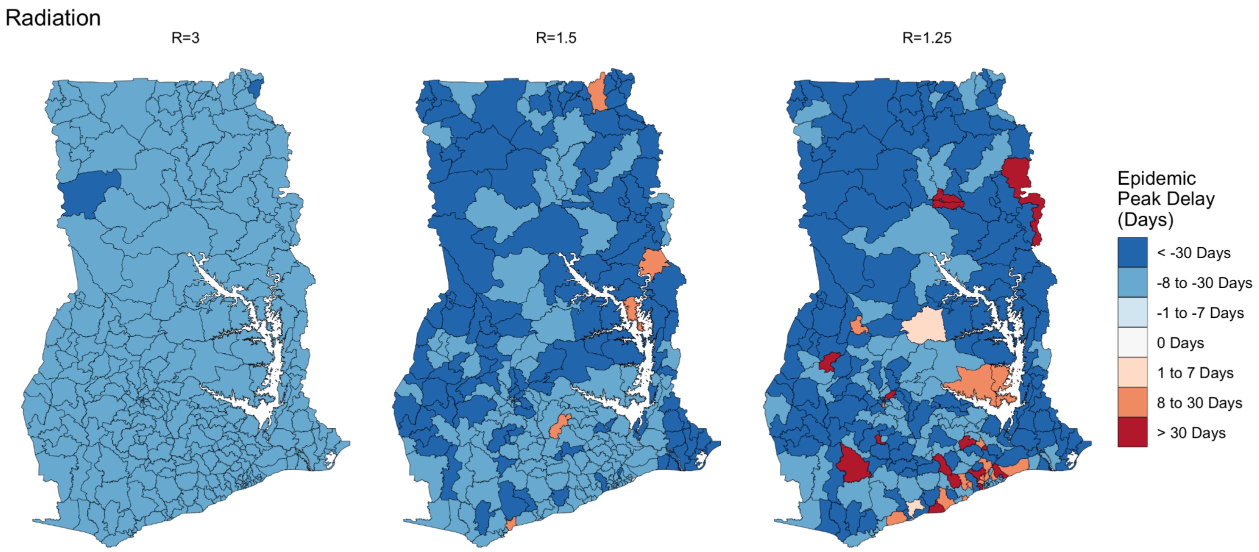


***Supplemental Figure 12. Influence of introduction location and $R_0$ on the difference between aggregation methodologies.*** *Difference between the timing of the peak of a modelled epidemic informed my mobility data modelled using the Radiation model. Negative numbers indicate that the epidemic predicted based on the all pairs methodology was earlier than the epidemic predicted based on the sequential methodology.*
